# Influence factors of death risk among COVID-19 patients in Wuhan, China: a hospital-based case-cohort study

**DOI:** 10.1101/2020.03.13.20035329

**Authors:** Lin Fu, Jun Fei, Hui-Xian Xiang, Ying Xiang, Zhu-Xia Tan, Meng-Die Li, Fang-Fang Liu, Hong-Yan Liu, Ling Zheng, Ying Li, Hui Zhao, De-Xiang Xu

## Abstract

**Background:** Coronavirus disease 2019 (COVID-19) triggered by infection with severe acute respiratory syndrome coronavirus-2 (SARS-CoV-2) has been widely pandemic all over the world. The aim of this study was to analyze the influence factors of death risk among 200 COVID-19 patients.

**Methods:** Two hundred patients with confirmed SARS-CoV-2 infection were recruited. Demographic data and clinical characteristics were collected from electronic medical records. Biochemical indexes on admission were measured and patient’s prognosis was tracked. The association of demographic data, clinical characteristics and biochemical indexes with death risk was analyzed.

**Results:** Of 200 COVID-19 patients, 163 (81.5%) had at least one of comorbidities, including diabetes, hypertension, hepatic disease, cardiac disease, chronic pulmonary disease and others. Among all patients, critical cases, defined as oxygenation index lower than 200, accounted for 26.2%. Severe cases, oxygenation index from 200 to 300, were 29.7%. Besides, common cases, oxygenation index higher than 300, accounted for 44.1%. At the end of follow-up, 34 (17%) were died on mean 10.9 day after hospitalization. Stratified analysis revealed that older ages, lower oxygenation index and comorbidities elevated death risk of COVID-19 patients. On admission, 85.5% COVID-19 patients were with at least one of extrapulmonary organ injuries. Univariable logistic regression showed that ALT and TBIL, two indexes of hepatic injury, AST, myoglobin and LDH, AST/ALT ratio, several markers of myocardial injury, creatinine, urea nitrogen and uric acid, three indexes of renal injury, were positively associated with death risk of COVID-19 patients. Multivariable logistic regression revealed that AST/ALT ratio, urea nitrogen, TBIL and LDH on admission were positively correlated with death risk of COVID-19 patients.

**Conclusion:** Older age, lower oxygenation index and comorbidities on admission elevate death risk of COVID-19 patients. AST/ALT ratio, urea nitrogen, TBIL and LDH on admission may be potential prognostic indicators. Early hospitalization is of great significance to prevent multiple organ damage and improve the survival of COVID-19 patients.

**Summary:** In this hospital-based case-cohort study, we found that serum urea nitrogen, TBIL, LDH and AST/ALT ratio, several markers of extrapulmonary organ injuries, were positively correlated with death risk of COVID-19 patients. We provide evidence for the first time that multiple organ damage on admission influences the prognosis of COVID-19 patients. Early hospitalization is beneficial for elevating the survival rate of COVID-19 patients especially critical ill patients.

## Introduction

Since December 2019, a cluster of acute respiratory illness patients with unclear causes have been found in some hospitals in Wuhan City, Hubei Province, China.^1^ On February12, 2020, the International Committee on Taxonomy of Viruses has declared that the official classification of the new coronavirus is severe acute respiratory syndrome coronavirus 2 (SARS-CoV-2). This is a novel member of human coronavirus, newly identified recently. World Health Organization (WHO) pronounces that the official name of the disease caused by the virus is corona virus disease 2019 (COVID-19). The main symptoms and signs of COVID-19 patients are with fever, dry cough, dyspnea, fatigue and lymphopenia. In severe and critical patients, SARS-CoV-2 infections caused viral pneumonia may lead to severe acute respiratory syndrome and even death. ^2-4^ After the first patient was reported in December 2019 in Wuhan City, Hubei Province, China, this infecting disease broke out quickly, spread around all over the world and was persistently evolving so far. ^5^ Until 6 March, 2020, 80581 cases were confirmed to have been infected with SARS-CoV-2 and 3016 cases died from SARS-CoV-2 infection in China. In other countries, total 17024 COVID-19 patients were confirmed and 343 patients died after being infected with SARS-CoV-2. ^6^

SARS-CoV-2 is mainly infected through the respiratory tract in humans. Several groups have reported the clinical characteristics of COVID-19 patients. Except for lung injury, infection with SARS-CoV-2 induces myocardial dysfunction, hepatic injury and even renal failure.^7^ The rate of overall case-fatality was about 2.3% across China. ^8^ Although few deaths were found in mild COVID-19 patients, the rate of case-fatality was obviously elevated among critical ill patients. Nevertheless, what factors influence the prognosis and death of COVID-19 patients have not been clarified.

The aim of the present study is to analyze risk factors of the prognosis and death among 200 COVID-19 patients. We show that older ages, lower oxygenation index and comorbidities are death risk factors of COVID-19 patients. We demonstrate for the first time that serum urea nitrogen, TBIL, LDH and AST/ALT ratio on admission may be potential prognostic indicators of COVID-19 patients.

## Methods

### Study Design and Participants

For this study, 220 patients with confirmed COVID-19 were recruited from Union Hospital of Huazhong University of Science and Technology from January 1 to January 30, 2020. All patients were clinically diagnosed with viral pneumonia on basis of typical clinical manifestations (fever or other respiratory symptoms) accompanied with the obvious chest radiology changes. Union Hospital of Huazhong University of Science and Technology is one of COVID-19-designated hospitals for the hospitalization of patients with COVID-19 in Wuhan City, Hubei Province, China. In order to respond to national summons positively, the Second Affiliated Hospital of Anhui Medical University enthusiastically sent to a medical team to recuse patients with COVID-19 in Wuhan City. All patients were initially detected with real-time RT-PCR assay for SARS-CoV-2 RNA, analyzed genetic sequence that matched COVID-19 and confirmed infection with SARS-CoV-2. Twenty patients with negative COVID-19 detection results were excluded in the present research. All 200 patients with COVID-19 were eligible in this research and we followed up the patients’ outcomes. Each COVID-19 patients gave advanced oral consent. This study was approved by the Ethics Committee of Anhui Medical University.

### Date collection

The medical records of each patients with COVID-19 were evaluated by research team in the Department of Respiratory and Critical Care Medicine, Union Hospital of Huazhong University of Science and Technology as well as the Second Affiliated Hospital of Anhui Medical University. Following fundamental data were collected from the electronic medical records of each patient: demographic information, preexisting comorbidities including chronic pulmonary disease, hepatic disease, cardiovascular disease, hypertension, diabetes and other disease, as well as exposure history, medical history, surgery history, signs and symptoms, chest computed tomographic (CT) scan and laboratory test results. The date of disease onset and hospital date, as well as death date were recorded. The onset time was defined as the date when patients’ any symptoms and signs were found.

### Laboratory testing

Patient’s pharyngeal swab specimens were collected for extracting SARS-CoV-2 RNA. Viral nucleic acid was detected via real-time reverse-transcriptase polymerase-chain-reaction (RT-PCR) assay with a COVID-19 nucleic acid detection kit following operating instructions (Shanghai bio-germ Medical Technology Co Ltd). All viral RNA extracting and nucleic acid detection for patients were executed by the Center for Disease Control and Prevention of Wuhan City. All medical laboratory tests, including white blood cell (WBC), neutrophil, lymphocyte, monocyte, basophil, total bilirubin (TBIL), direct bilirubin (DBIL), alanine aminotransferase (ALT), aspartate aminotransferase (AST), creatinine, urea nitrogen, uric acid, creatine kinase, creatine kinase isoenzyme, oxygenation index were examined on admission. The definition of organ injuries was that any of the biochemical indexes of liver function, renal function or myocardial function were beyond normal range. ^9^ All medical laboratory tests were analyzed by the clinical laboratory of Union Hospital of Huazhong University of Science and Technology.

### Statistical analysis

All statistical analyses were performed using SPSS 21.0 software. Categorical variables were expressed with frequencies and percentages. Continuous variables were shown using median and mean values. Means for continuous variables were compared with independent-samples *t* tests when the data were normally distributed; if not, the Mann-Whitney test was used. Proportions for categorical variables were compared with the *chi-square* and *Fisher’s* exact test. Univariable logistic regression between basic disease or different parameter and demise was performed. Moreover, the main risks related with demise were examined using multivariable logistic regression models adjusted for potential confounders. Statistical significance was determined at *P*<0.05.

## Results

### 1. Demographics data and clinical characteristics

The clinical characteristics of all patients are descripted in Table 1. Of 200 cases, 99 (49.3%) patients were males. 49 (24.5%) cases were younger than 49 years, 53 (26.5%) cases between 50 to 59 years, 59 (29.5%) cases between 60 to 69 years, and 39 (19.8%) cases older than 70 years. Among all cases, 163 (81.5%) had at least one of comorbidities, such as diabetes (68.2%), hypertension (51.5%), hepatic disease (4.5%), cardiac disease (8.0%), chronic pulmonary disease (4.0%) and others (3.5%). The most common symptom of COVID-19 patients was fever (88.0%), followed by diarrhea (59.7%), fatigue (52.2%) and cough (46.3%) (Supplemental Table 1). Among all patients, critical cases, defined as oxygenation index lower than 200, accounted for 51 (26.2%). Severe cases, oxygenation index from 200 to 300, were 58(29.7%). Besides, common cases, oxygenation index higher than 300, accounted for 88(44.1%). The median oxygenation index was 282.9, far below normal range. The blood routine analysis showed that median WBC count was 5.05×10 ^9^/L. As shown in Supplemental Table 2, median neutrophil count was 3.51×10 ^9^/L and median lymphocyte count was 0.88×10^9^/L.

**Table 1.** Clinical characteristics and comorbidities of COVID-19 patients.

| Characteristics | Total cases (N) | Dead cases (N, %) | RR (95% CI) | P |
| --- | --- | --- | --- | --- |
| **Gender** |  |  |  |  |
| Male | 99 | 16 (16.2) | 0.907 (0.491, 1.675 ) | 0.755 |
| Female | 101 | 18 (17.8) | 1 | — |
| **Age** |  |  |  |  |
| <49 year | 49 | 2 (4.08) | 1 | — |
| 50-59 | 53 | 8 (15.1) | 3.698 (0.825, 16.574) | 0.062 |
| 60-69 | 59 | 7 (11.9) | 2.907 (0.632, 13.359) | 0.145 |
| >70 | 39 | 17 (43.6) | 10.679 (2.624, 43.459) | <0.001 |
| **Smoker<sup>a</sup>** |  |  |  |  |
| Yes | 89 | 16 (16.2) | 0.809 (0.443,1.478) | 0.490 |
| No | 81 | 18 (17.8) | 1 | — |
| **Oxygenation index** |  |  |  |  |
| <200 | 51 | 27 (52.9) | 15.176 (4.847, 47.519) | <0.001 |
| 200-300 | 58 | 4 (6.90) | 4.095 (0.975, 17.197) | 0.039 |
| >300 | 86 | 3 (3.49) | 1 | — |
| **Comorbidities** |  |  |  |  |
| Yes | 161 | 37 (23.0) | 2.969 (0.965, 9.131) | 0.033 |
| No | 39 | 3 (7.69) | 1 | — |
| **Diabetes** |  |  |  |  |
| Yes | 137 | 26 (19.0) | 1.495 (0.717, 3.114) | 0.272 |
| No | 63 | 8 (12.7) | 1 | — |
| **Hypertension** |  |  |  |  |
| Yes | 101 | 22 (21.8) | 1.797 (0.941, 3.430) | 0.069 |
| No | 99 | 12 (12.1) | 1 | — |
| **Hepatic diseases** |  |  |  |  |
| Yes | 9 | 2 (22.2) | 1.326 (0.375, 4.688) | 0.670 |
| No | 191 | 32 (16.8) | 1 | — |
| **Cardiac disease** |  |  |  |  |
| Yes | 16 | 2 (12.5) | 0.719 (0.189, 2.729) | 0.617 |
| No | 184 | 32 (17.4) | 1 | — |
| **Pulmonary disease** |  |  |  |  |
| Yes | 8 | 4 (50.0) | 3.200 (1.486, 6.890) | 0.011 |
| No | 192 | 30 (15.6) | 1 | — |
| **Other disease** |  |  |  |  |
| Yes | 7 | 2 (28.6) | 1.723 (0.512, 5.798) | 0.407 |
| No | 193 | 32 (16.6) | 1 | — |

a: Information of 56 smokers was missing.

**Table 2.** Biochemical indexes on admission to hospital of COVID-19 patients.

| Parameter | Normal range | Total cases<br>(N=200) | Alive cases<br>(N=166) | Dead cases<br>(N=34) | *P* |
| --- | --- | --- | --- | --- | --- |
| **Liver function indexes** |  |  |  |  |  |
| Total bilirubin (μm/L) | 5.1-20.5 | 11.4 (9.0, 15.6) | 10.9 (8.75, 15.1) | 14.8 (11.6, 18.3) | 0.007 |
| Direct Bilirubin (μm/L) | 0-6.8 | 3.3 (2.3, 5.0) | 3.10 (2.20, 4.33) | 5.05 (3.78, 6.48) | 0.007 |
| Alanine aminotransferase (U/L) | 5-40 | 23.5 (16.0, 42.0) | 22.0 (16.0, 41.0) | 34.5 (17.8, 55.8) | 0.030 |
| **Renal function indexes** |  |  |  |  |  |
| Creatinine (μm/L) | 44-115 | 66.0 (54.0, 87.0) | 65.0 (53.5, 83.0) | 90.5 (61.5, 113.5) | <0.001 |
| Urea nitrogen (mm/L) | 1.7-7.1 | 4.5 (3.3, 6.4) | 4.1 (3.2, 5.7) | 7.40 (5.9, 11.1) | <0.001 |
| Uric acid (μm/L) | 208-428 | 257.0 (190.0, 323.0) | 244.0 (189.3, 307.3) | 280.0 (229.8, 430.8) | 0.009 |
| **Myocardial function indexes** |  |  |  |  |  |
| Creatine kinase (U/L) | 38-174 | 90.5 (53.8, 172.5) | 79.5 (50.0, 144.5) | 180.5 (80.8, 404.5) | <0.001 |
| Creatine kinase isoenzymes (N) | Negative | 150 (74.6%) | 128 (77.1%) | 22 (64.7%) | 0.147 |
| Myoglobin (N) | Negative | 119 (59.2%) | 106 (63.9%) | 13 (38.2%) | 0.001 |
| Cardiac troponin I (N) | Negative | 137 (68.5 %) | 112 (67.5 %) | 25 (73.5 %) | 0.770 |
| Lactate dehydrogenase (U/L) | 109-245 | 274.0 (190.0, 403.0) | 245.0 (179.0, 345.0) | 495.5 (350.8, 695.8) | <0.001 |
| Aspartate aminotransferase (U/L) | 8-37 | 35.0 (23.4, 56.8) | 32.0 (22.8, 46.3) | 61.5 (40.3, 98.5) | 0.000 |
| Aspartate aminotransferase/ Alanine aminotransferase ratio | 0.14~5 | 1.43 (1.07, 1.94) | 1.37 (1.03, 1.77) | 2.00 (1.34, 2.74) | 0.000 |

### 2. Association of demographic data, clinical characteristics and comorbidities with death risk of COVID-19 patients

Thirty-four patients were died on mean 10.9 day after hospitalization. The association of demographic data with death risk of COVID-19 patients was analyzed. As shown in Table 1, fatality rate was 16.2% in male patients and 17.8% in female patients. Moreover, no difference on the fatality rate of COVID-19 patients was found between smokers and nonsmokers. The influence of ages on the fatality rate of COVID-19 patients is presented in Table 1. The fatality rate was 4.08% in COVID-19 patients younger than 40 years old, 8 (15.1%) patients between 50 and 59 years old, 7 (11.9%) patients between 60 and 69 years old, and 17 (43.6%) patients over 70 years old. The *RR* was 3.698 (95% *Cl*: 0.825, 16.574; *P*=0.062) in COVID-19 patients between 50 and 59 years old, 2.907 (95% *Cl*: 0.632, 13.359; *P*=0.145) in patients between 60 and 69 years old, and 10.679 (95% *Cl*: 2.624, 43.459; *P*<0.001) in patients over 70 years old, respectively. The relationship between oxygenation index and death risk of COVID-19 patients was analyzed. As shown in Table 1, the fatality rate was 52.9% in critical ill cases, 6.90% in severe cases, and 3.49% in common cases, respectively. The *RR* was 15.176 (95% *Cl*: 4.847, 47.519; *P*<0.001) in critical ill cases with COVID-19 and 4.095 (95% *Cl*: 0.975, 17.1979; *P*=0.039) in severe cases, respectively. The correlation between comorbidities and death risk of COVID-19 patients was then evaluated. As shown in Table 1, the fatality rate was 23.0% in COVID-19 patients with at least one of comorbidities, remarkably higher than 7.69% in COVID-19 patients without comorbidity. The *RR* was 2.969 (95% *Cl*: 0.965, 9.131; *P*=0.033) in COVID-19 patients with comorbidities. Further analysis found that the fatality rate was 50% in COVID-19 patients with chronic pulmonary disease, remarkably higher than 15.6% in COVID-19 cases without chronic pulmonary disease. The *RR* was 3.200 (95% *Cl*: 1.486, 6.890; *P*=0.011) in subjects with chronic pulmonary disease. No statistically significant association was observed between death risk and other coexisting comorbidities, such as diabetes, hypertension, hepatic disease, cardiac disease and other chronic diseases (Table 1).

### 3. Associations between biochemical indexes and death risk of COVID-19 patients

The present study found that 182 COVID-19 patients (85.5%) were with at least one of extrapulmonary organ injuries, including 66 (33.0%) with liver injury, 45 (22.5%) with acute kidney injury, 148 (74.0%) with cardiac injury. The association between indexes of hepatic injury on admission and death risk was analyzed among 200 COVID-19 patients. As shown in Table 2, median TBIL was 11.4 μm/L, whereas median DBIL was 2.3 μm/L (Table 2). TBIL of 17 (8.5%) patients and DBIL of 19 (9.5%) patients were beyond normal range. The median ALT was 23.5 U/L. ALT of 56 (28.6%) patients were above 80 U/L (Table 2). Univariable logistic regression analysis showed that serum ALT (*OR*=1.403; 95% *Cl*: 1.020,1.929; *P*<0.05) and TBIL (*OR*=12.113; 95% *Cl*: 1.905, 77.014; *P*<0.01) were positively with death risk of COVID-19 patients. There was no remarkable association between serum DBIL and death risk of COVID-19 patients (Table 3). The relationship between renal function markers on admission and death risk was evaluated. The results revealed that median creatinine was 66.0 μm/L among COVID-19 patients, of which 17 (8.5%) cases were above normal range. The median urea nitrogen was 4.5 mm/L among COVID-19 patients, of which 36 cases (18.0%) were beyond normal range. The median uric acid was 257.0 μm/L and 17 cases (8.5%) were above normal limits. Univariable logistic regression analysis indicated that serum creatinine (*OR*=2.094; 95% *Cl*: 1.413, 3.102; *P* < 0.001), urea nitrogen (*OR*=4.041; 95% *Cl*: 2.344, 6.967; *P*<0.001) and uric acid (*OR*=1.794; 95% *Cl*: 1.265, 2.546; *P*<0.001) were positively with death risk of COVID-19 patients (Table 3). Finally, the correlation between myocardial enzyme parameters and death risk was revealed in Table 3. The median LDH, creatine kinase and AST were 274.0, 90.5, and 35.0 U/L, respectively. Further analysis showed that serum AST and LDH of half patients were far beyond normal range. As shown in Table 2, AST/ALT ratio was 1.43 among 200 patients. The number of serum myoglobin-positive patients was 119 (59.2%), whereas the number of serum cardiac troponin I-positive patients was 137 (68.5%). As shown in Table 3, univariable logistic regression analysis found that there was a positive correlation between serum creatine kinase (*OR*=2.127; 95% *Cl*: 1.439, 3.147; *P*<0.001), myoglobin (*OR*=3.624; 95% *Cl*: 1.590, 8.259; *P*<0.001), LDH (*OR*=5.929; 95% *Cl*: 3.116, 11.281; *P*<0.001), AST (*OR*=2.627; 95% *Cl*: 1.586, 4.351; *P*<0.001), and AST/ALT ratio (*OR*=3.279; 95% *Cl*: 1.917, 5.607; *P*<0.001) with death risk of COVID-19 patients (Table 3). Multivariable logistic regression was used to analyze the relationship between biochemical indexes and death risk of COVID-19 patients after adjustment for potential confounding factors. The results found that only serum TBIL (*OR*=1.062; 95% *Cl*: 1.007, 1.120; *P*<0.05), urea nitrogen (*OR*=1.589; 95% *Cl*: 1.273, 1.984; *P*<0.001), LDH (*OR*=10.395; 95% *Cl*: 2.163, 49.957; *P*<0.01) and AST/ALT ratio (*OR*=3.224; 95% *Cl*: 1.586, 6.555; *P*<0.001) were positively associated with death risk of COVID-19 patients. However, there was no relationship between other biochemical indexes and death risk of COVID-19 patients (Table 4).

**Table 3.** Univariable logistic regression between biochemical indexes and death risk of COVID-19 patients.

| Parameter | $\beta$ | Wald | P | OR (95% CI) |
| --- | --- | --- | --- | --- |
| **Liver function indexes** |  |  |  |  |
| Total bilirubin | 2.494 | 6.985 | 0.008 | 12.113 (1.905, 77.014) |
| Direct Bilirubin | 0.932 | 2.167 | 0.141 | 2.539 (0.734, 8.781) |
| Alanine aminotransferase | 0.338 | 4.335 | 0.037 | 1.403 (1.020, 1.929) |
| **Renal function indexes** |  |  |  |  |
| Creatinine | 0.739 | 13.583 | 0.000 | 2.094 (1.413, 3.102) |
| Urea nitrogen | 1.396 | 25.248 | 0.000 | 4.041 (2.344, 6.967) |
| Uric acid | 0.585 | 10.728 | 0.001 | 1.794 (1.265, 2.546) |
| **Myocardial function indexes** |  |  |  |  |
| Creatine kinase | 0.755 | 14.323 | 0.000 | 2.127 (1.439, 3.147) |
| Creatine kinase isoenzymes | 0.711 | 2.055 | 0.152 | 2.038 (0.770, 5.384) |
| Myoglobin | 1.288 | 9.386 | 0.002 | 3.624 (1.590, 8.259) |
| Cardiac troponin I | -0.446 | 0.463 | 0.496 | 0.640 (0.177, 2.314) |
| Lactate dehydrogenase | 1.780 | 29.412 | 0.000 | 5.929 (3.116, 11.281) |
| Aspartate aminotransferase | 0.966 | 14.078 | 0.000 | 2.627 (1.586, 4.351) |
| Aspartate aminotransferase/ Alanine aminotransferase ratio | 1.187 | 18.816 | 0.000 | 3.279 (1.917, 5.607) |

**Table 4.** Multivariable logistic regression between biochemical indexes and death risk of COVID-19 patients.

| Parameter | $\beta$ | Wald | *P* | OR (95% CI) |
| --- | --- | --- | --- | --- |
| **Liver function indexes** |  |  |  |  |
| Alanine aminotransferase | 0.006 | 1.112 | 0.292 | 1.006 (0.955, 1.017) |
| Total bilirubin | 0.060 | 4.980 | 0.026 | 1.062 (1.007, 1.120) |
| **Renal function** |  |  |  |  |
| Creatinine | -0.011 | 0.112 | 0.191 | 0.989 (0.973, 1.005) |
| Urea nitrogen | 0.463 | 16.731 | <0.001 | 1.589 (1.273, 1.984) |
| Uric acid | 0.000 | 0.112 | 0.738 | 0.999 (0.994, 1.004) |
| **Myocardial function** |  |  |  |  |
| Creatine kinase | 0.000 | 0.126 | 0.723 | 1.000 (0.999, 1.001) |
| Myoglobin | -0.441 | 0.692 | 0.405 | 0.643 (0.227, 1.819) |
| Lactate dehydrogenase | 2.341 | 8.545 | 0.003 | 10.395 (2.163, 49.957) |
| Aspartate aminotransferase | -0.001 | 0.032 | 0.858 | 0.999 (0.999, 1.001) |
| Aspartate aminotransferase/<br>Alanine aminotransferase ratio | 1.171 | 10.460 | 0.001 | 3.224 (1.586, 6.555) |

## Discussion

The present study aimed to analyze influence factors of death risk among 200 COVID-19 patients. The main findings of the present study include: (1) older age and comorbidities are risk factors of death in COVID-19 patients; (2) lower oxygenation index on admission elevates death risk of COVID-19 patients. (3) serum urea nitrogen, TBIL, LDH and AST/ALT ratio on admission are potential indicators of death risk among COVID-19 patients.

Up to March 6, 2020, SARS-CoV-2 infection has caused 3016 deaths all over China. ^10^ As no specific drug, it is important to clarify which factors increase death risk of COVID-19 patients. In the present study, we analyzed the association between major demographic characteristics and death risk of COVID-19 patients. Our results showed that no significant difference on the fatality rate was observed between males and females. A recent study indicated that smoking increases the risk of infection with SARS-CoV-2. ^11^ In the present study, we analyzed the influence of smoking on death risk of COVID-19 patients. Unexpectedly, there was no difference on the fatality rate between smokers and nonsmokers. A recent report showed that elderly patients with critical COVID-19 were more likely to die. ^12^ The present study found that the fatality rate was 4.08% in COVID-19 patients younger than 40 years old, 15.1% in patients between 50 and 59 years old, 11.9% in patients between 60 and 69 years old, and 43.6% in patients over 70 years old. Our results provide additional evidence that older age elevates death risk of COVID-19 patients.

Several reports demonstrated that COVID-19 patients with comorbidities were at an increased death risk. ^2^ Indeed, the present study showed that 161 COVID-19 patients (80.5%) were with at least one of comorbidities, including diabetes, hypertension, hepatic disease, cardiac disease, chronic pulmonary disease and others. Thus, we analyzed the association between comorbidities and death risk of COVID-19 patients. Our results showed that comorbidities obviously elevated death risk of COVID-19 patients. Further analysis found that neither cardiac nor hepatic diseases were associated with death risk of COVID-19 patients. Interestingly, COVID-19 patients with chronic pulmonary disease, mainly COPD, obviously elevated death risk. The present study showed that more than half COVID-19 patients were with either diabetes or hypertension. Therefore, it is especially interesting whether these comorbidities increase death risk of COVID-19 patients. We found that there was a trend for hypertension and diabetes to elevate the risk of death from COVID-19 pneumonia. These results need to be further demonstrated in a larger sample investigation.

Previous studies found that critical ill patients are more likely to die from COVID-19 pneumonia. ^8, 12^ Among 200 COVID-19 patients, critical ill cases, defined as oxygenation index lower than 200, accounted for 26.2%. Severe cases, oxygenation index from 200 to 300, were 29.7%. In addition, common cases, whose oxygenation index higher than 300, accounted for 44.1%. The present study analyzed the influence of admission oxygenation index on death risk of COVID-19 patients. We showed that fatality rate in critical ill cases, whose oxygenation index was lower than 200, was 52.9%, significantly higher than 6.90% in severe cases and 3.49% in common cases. These results suggest that lower oxygenation index on admission elevates death risk of COVID-19 patients.

Accumulating data demonstrate that angiotension converting enzyme (ACE)2, as a receptor for SARS-CoV-2, is abundantly expressed in airway epithelial cells and plays a crucial role in the pathogenesis of SARS-CoV-2 infection. ^13^ Recently, several groups reported that ACE2 was expressed in renal tubular cells, hepatic cholangiocytes and cardiocytes. ^14-16^ Indeed, multiple organ damage is usually accompanied at the early stage of COVID-19 pneumonia. ^17, 18^ The present study showed that 85.5% COVID-19 patients were with at least one of extrapulm74.0% with cardiac injury. Thus, univariable logistic regression was used to analyze the association between biochemical indexes of extrapulmonary organ injury on admission and death risk of COVID-19 patients. As expected, serum ALT and TBIL levels, two markers of liver injury, were positively associated with fatality rate of COVID-19 patients. Moreover, creatinine, urea nitrogen and uric acid, three serum indexes of kidney injury, had a prominently positive association with death risk of COVID-19 patients. In addition, the increased creatine kinase, myoglobin, LDH, AST and AST/ALT ratio, several serum markers of cardiac injury, elevated death risk of COVID-19 patients. To exclude potential confounding factors, multivariable logistic regression was used to further analyze the relationship between biochemical indexes of extrapulmonary organ injuries and death risk of COVID-19 patients. We found that serum urea nitrogen, TBIL, LDH and AST/ALT ratio on admission were positively correlated with death risk of COVID-19 patients after adjustment for potential confounding factors. These results suggest that extrapulmonary organ injuries on admission may be potential indicators of death risk among COVID-19 patients. Therefore, early hospitalization is of great significance to prevent multiple organ damage and improve the survival of COVID-19 patients.

In summary, the present study analyzed influence factors of prognosis and death among 200 COVID-19 patients. We showed that older ages, lower oxygenation index and comorbidities on admission elevated death risk of COVID-19 patients. Serum urea nitrogen, LDH, TBIL and AST/ALT ratio on admission are potential indicators of death risk among COVID-19 patients. We provide evidence for the first time that multiple organ damage on admission may influence prognosis of COVID-19 patients. Early hospitalization is beneficial for elevating the survival rate of COVID-19 patients especially critical ill patients.

## Data Availability

We declare that all data referred to in the manuscript is availability.

## Note

## Acknowledgments

We thank all patients and their families involved in this research. We also thank all members of respiratory and critical care medicine in the Second Affiliated Hospital of Anhui Medical University and Union Hospital of Huazhong University of Science and Technology for recruiting participators.

## Contributors

DXX, HZ, LF and JF designed research; LF, JF, HXX, YX, ZXT, MDL, FFL, HYL, LZ and YL conducted research; LF analyzed data; DXX and LF wrote the paper; DXX and LF had primary responsibility for final content. All authors read and approved the final manuscript.

## Disclosure Statement

The authors have nothing to disclose.

## Funding

This study was supported by National Natural Science Foundation of China (81630084) and National Natural Science Foundation Incubation Program of the Second Affiliated Hospital of Anhui Medical University (2019GQFY06).

## Conflicts of interest

Lin Fu, Jun Fei, Hui-Xian Xiang, Ying Xiang, Zhu-Xia Tan, Meng-Die Li, Fang-Fang Liu, Hong-Yan Liu, Ling Zheng, Ying Li, Hui Zhao and De-Xiang Xu declared that there were no competing interests.

## Notes

### Competing Interest Statement

The authors have declared no competing interest.

### Clinical Trial

Coronavirus disease 2019 (COVID-19) triggered by infection with severe acute respiratory syndrome coronavirus-2 (SARS-CoV-2) has been widely pandemic all over the world. After the first patient was reported in December 2019 in Wuhan City, Hubei Province, China, this infecting disease broke out quickly, spread around all over the world and was persistently evolving so far. Until 6 March, 2020, 80581 cases were confirmed to have been infected with SARS-CoV-2 and 3016 cases died from SARS-CoV-2 infection in China. In other countries, total 17024 COVID-19 patients were confirmed and 343 patients died after being infected with SARS-CoV-2. This novel disease is a neo-type respiratory contagious disease that has high infectivity and mortality as well as heavy damage on public health. Clinical trial has been registered before this research. However, it needs a long time to finish all the process, the clinical trial ID is not obtained temporarily and still transacted.

